# Public knowledge and attitude towards epilepsy and its associated factors in Debre Berhan, North Shoa, Amhara Region, Ethiopia, 2018/19. Community-based cross-sectional study

**DOI:** 10.1101/19001578

**Authors:** Abate Dargie, Nigus Alemnew, Elyas Admasu

## Abstract

**Introduction:** Epilepsy is chronic brain disorder characterized by recurrent derangement of the nervous system due to the sudden excessive disorderly discharge of the cerebral neurons. People living with epilepsy continue to suffer from enacted or perceived stigma that is based on myths, misconceptions, and misunderstandings that have persisted for many years. Therefore, this study aimed to assess the community general knowledge and attitude towards epilepsy.

**Methods:** Community-based cross-sectional study was conducted to assess public general knowledge and attitude towards epilepsy and its associated factors using structured pretested questionnaire. Data were entered into Epi data version 3.1 and transported to SPSS version 21 further analysis. Both Bivariable and Multivariable Logistic Regression was done to identify associated factors. Odds Ratios and their 95% Confidence interval were computed and variables with p-value less than 0.05 was considered significantly associated factors.

**Results:** 596 study participants participated in a response rate of 98%. Among the study participants, 43.6 (95% CI: 39.6, 47.5) had poor knowledge and 41.3 (95% CI: 37.4, 45.1) had an unfavorable attitude. Being secondary education, marital status, witnessed a seizure and heard the term epilepsy were showed statistically significant association with poor knowledge about epilepsy. Level of education, low average monthly income, not witnessed a seizure, not heard the term epilepsy and distant from health facility showed statically significant association with the unfavorable attitude.

**Conclusion:** In this study, Debre Berhan communities were found to have deficits in terms of general knowledge and attitude about epilepsy; and it should be given due attention.

## 1. Introduction

Epilepsy is a chronic brain disorder characterized by recurrent derangement of the nervous system due to the sudden excessive disorderly discharge of the cerebral neurons that results in an almost instantaneous disturbance of sensation and loss of consciousness(1). It is becoming a burden for more than 70 million people in the world. Nearly 80% of the people with epilepsy are found in developing countries, where the disease remains a major public health problem, not only because of its health implications but also for its social, cultural, psychological and economic effects (2, 3). Epilepsy contributed more than 7 million to the global burden of diseases in 2000 measured in disability-adjusted life years (DALYs). It is apparent that close to 90% of the worldwide burden of epilepsy is found in developing regions, with more than half occurring in the 39% of the global population living in countries with the highest levels of premature mortality, the lowest levels of premature mortality and the least levels of income(4, 5).

The reported prevalence of active epilepsy in developing countries ranges from 5 to 10 per 1,000 people, and it is more prevalent in rural communities(6, 7). Universally and all over history, epilepsy has been culturally an undervalued condition(8). Epileptic patients and their families have suffered isolation by society and deprivation of treatment, leading to frequent injuries and death (9). Epilepsy is a fascinating topic by nature and is poorly understood by the public, even among people who know someone with the disorder. Lack of knowledge about the causes of epilepsy has been associated with negative attitudes, beliefs, and stigma (3, 6, 10). Lack of understanding about epilepsy is a leading cause of discrimination in the workplace and in schools (11).

Most studies showed that a high number of people ever heard about epilepsy. However, most of them had a lower level of knowledge. A significant number of people argued that epilepsy is a mental disease, hereditary disease, contagious, evil spirit or God’s curse (12, 13). Due to the negative attitude towards epilepsy, many people do not want to work or live with epileptics. Large numbers of people do not want even to shake hands with epileptic individuals, and they try to keep their children away from these patients (14).

The Causes and risk factors of epilepsy include head injury, perinatal cause, infectious disease, brain tumor, cerebrovascular disease, febrile convulsions and familial past history of epilepsy The annual incidences and prevalence of epilepsy in sub-Saharan Africa is high (63–158 per100 000 inhabitants) and 15 per 1000 people respectively. The mean sex ratio of men to women is 1:4 in studies from sub-Saharan Africa(15).

The incidence of epilepsy in Ethiopia is 64 per 100,000. The age-specific incidence was highest in the group aged 0-9 years; the next highest incidence was that in the group aged 10-19 years; the third highest was in the group aged 20-29 years(16).

People living with epilepsy continue to suffer from enacted or perceived stigma that is based on myths, misconceptions, and misunderstandings that have persisted for many years. In the last decade, there has been an increase in individual literacy rate and increased access to technology in Debre Berhan population. However, it is unclear if this has any effect on knowledge, attitude, and practice (KAP) attitude toward epilepsy.

Fear, misunderstanding, lack of awareness, wrong beliefs and the resulting social stigma and discrimination compounding epilepsy make epileptic patients live poor quality life(17).

Sociocultural attitudes continue to cause a negative impact on the management of epilepsy in many African nations. The disorder is enrobed in superstition, discrimination, and stigma in many of these states. Religious and sociocultural beliefs influence the nature of treatment and maintenance received by people with epilepsy. Many communities in Africa and other developing countries believe that epilepsy results from witchcraft or possession by evil spirits, and hence, treatment should be through the use of herbs from traditional doctors, fetish priests, and religious leaders. Persons with epilepsy are rejected and discriminated against in education, employment, and marriage in Africa because epilepsy is seen as a highly contagious and shameful disease in the eyes of the public(18).

A compiled data of articles across the world showed that the magnitude of poor knowledge towards epilepsy ranges from 40% to 86 % (19-23). Unfavorable attitude also ranges from 28% to 87% (11, 21-26) Studying knowledge and attitude towards epilepsy in Debre Berhan town enables to know the level of poor knowledge and unfavorable attitude and also it enables to dig out the factors that result in poor knowledge and unfavorable attitude.

Having poor knowledge and unfavorable attitude results in stigmatization and this stigmatization results in delay in medical treatment or medication adherence problems.

Findings could benefit policymakers, the district health team, community members, affected families, and people with epilepsy in designing strategies. Furthermore, the study possibly generates information in the area of the topic for large scale researchers to investigate further empirical evidence to control those factors attributable to poor knowledge and unfavorable attitude towards epilepsy in the study area.

This study aimed at studying the magnitude of knowledge and attitude of Debre Berhan community towards epilepsy and associated factors for poor community’s knowledge and unfavorable community’s attitude.

## 2. Methods

The study was conducted at Debre Berhan; the largest town of North Shewa of Amhara region which is located 130 km northeast of Addis Ababa from February 28-March 1, 2019. The total sample size of the study was 596 with a response rate of 98%. The source population was all households in Debre Berhan town. The study population was all households found in the selected Kebele. Each selected Household’s and avails during the data collection period were sampling units of the study and each individual in the selected household in which the actual data is collected were study units. Individuals who are severely ill to give the desired information were excluded from the study. Authors had selected 2 Kebele (22%) out of the total 9 Kebele using lottery method. Then using systematic random sampling method households for our study from the selected Kebele were selected by using a sampling fraction 8 and our starting household selected by using lottery method from the first 8 households, since we don’t have sampling frame we gave a code for every house. For those households that had more than one eligible participants, 1 study unit was selected by using the lottery method.

### 2.1. Study variables and operational definitions

The data was collected by using pretested 10 and 12 item screening tools adopted from related studies for knowledge and attitude respectively. The data collectors have interviewed the study participants in the households that can address the common socio-demographic, information and experience, and health facility related characteristics and questions that can assess the community’s knowledge and attitude towards epilepsy. The data were collected by five trained undergraduate degree students. The questioner was translated from English to Amharic and back to English to check the consistency. To ensure quality, data were checked for completeness, accuracy, clarity, and consistency by the principal investigator. To maintain the quality of the data the questionnaire was pre-tested among 5% (30 individuals) of the study households, and the pretested data was not included in data analysis. Intensive training for data collectors and Close supervision was held during the whole data collection period by the research advisor. The first draft of the questionnaire was evaluated by our advisers. Every morning of the study period, Orientation was given to members of data collectors and supervisors about how to take timely action to minimize errors. The reliability of the checklist was cheeked by Cronbach’s test and it was 0.9 for attitude and 0.81 for knowledge screening tools.

#### Independent variables

Socio-demographic factors: - age, sex, religion, marital status, address, academic status, residence, family role, ethnicity.

Health facility factor: - availability, distance

Experience with epilepsy: - having a family member with epilepsy, having epileptic friend, witnessed a seizure

Information factors: - magazines, TV, radios, family members, health care provider, training

#### Dependent variables

Level of Knowledge and Attitude towards epilepsy

#### Attitude

The section on attitude had 12 items and four Likert scale response options consisting of ‘strongly agree, agree, disagree and strongly disagree’. The response options were given (scored) a point of 4, 3, 2, and 1 respectively. The minimum score is 12 and the maximum score is 48. Individuals who scored below the mean (x□ =34) and ≥ mean (x□ =34) were categorized as unfavorable and favorable attitude respectively.

#### Knowledge

the general knowledge of the respondents about epilepsy was assessed by using 10 item checklist which has “Yes” /No choices. For each correct response to questions on knowledge, one point was given and “Zero” point was given for wrong answers. Based on this grading, a total of 10 points was allocated to the section on knowledge about epilepsy, such that those who scored 7–10 points were considered as having Good knowledge, 0-6 points had poor knowledge.

For internal consistency (reliability) of the knowledge and attitude instrument, Cronbach Alpha was used and its value was 0.808 and 0.90 respectively.

#### Operational definitions

##### Knowledge about epilepsy

individuals who scored above and below the mean score based on 10 item knowledge tool towards epilepsy was considered as good and poor knowledge respectively.

##### Attitude towards epilepsy

individuals who scored above and below the mean score based on 12 item attitude tool towards epilepsy was considered as favorable and unfavorable attitude respectively.

### 2.2. Data analysis

After the data, checked for completeness, and consistencies, then coded, entered, using Epi-data version 3.1 and exported to SPSS version 21 for analysis. Descriptive statistics were analyzed and presented by using tables, charts, and figures. Both Bivariable and multivariable Binary logistic regression was conducted to identify the associated factors with the outcome variables. Variables with a p-value less than 0.2 during Bivariable analysis was exported to multivariable analysis to control confounding effect. Variables with a p-value less than 0.05 and odds ratio with their 95% confidence interval during multivariable analysis were considered as significantly associated factors with the outcome variables. Assumption tests and model goodness of fit was checked and Hosmer Lemeshow test p-value of 0.79.

## 3. Results

### 3.1 Socio-demographic characteristics

A total of 606 households were selected for the sample, and 596 were successfully interviewed, yielding a response rate of 98%. Among the study participants, 332 (55.7%) were males and 264 (44.3%) were females. The median age of the respondents was 30 years and the interquartile range was 15 years. Among the 596 study participant’s majority 470(78.9%) were orthodox religion followers followed by 81(13.6%) Muslim and 45 (7.6%) protestant. Majority of the respondent 494(82.9%) were Amhara ethnicity followed by Oromo which accounts for 67(11.2%). Regarding the income of the respondents near to half 270 (45.3%) had a monthly income of >3000 followed by those whose monthly income is between 1000 & 2999 and < 1000 which accounts 243 (40.8%) and 83 (13.9%) respectively. Among the total participants, more than half 333 (55.9%) were married whereas the other 263 (44.1%) were single. Majority 445 (74.7%) of the respondent had a family size 1-3 followed by those whose family members were 4-5 136 (22.8%) (See table 1).

**Table 1.** Socio-demographic characteristics of the study participants, in DBT, Ethiopia, 2019

| Variables |  | Frequency | Percent |
| --- | --- | --- | --- |
| Age in years | 18-35 | 372 | 62.4 |
|  | 35-45 | 130 | 21.8 |
|  | >45 | 94 | 15.8 |
| Occupation | A farmer or housewife | 98 | 16.4 |
|  | Private workers | 170 | 28.5 |
|  | Daily laborer | 61 | 10.2 |
|  | Student | 65 | 10.9 |
|  | Government employee | 202 | 33.9 |
| Educational status | Not attend modern education | 116 | 19.5 |
|  | Primary | 95 | 15.9 |
|  | Secondary & pre | 145 | 24.3 |
|  | >12 grade | 240 | 40.3 |
| Family role | Family head | 386 | 64.8 |
|  | Son/daughter | 91 | 15.3 |
|  | Live alone | 119 | 20 |

### 3.2. Information, Experience and Health facility-related factors

The majority of the respondents 546 (91.6%) had heard of epilepsy as a disorder. The commonest sources of information about epilepsy were from their family or friends 305(55.9%) while the rest were through radio/magazines, TV/training and health professionals which account 92 (16.8%), 85 (15.6%) and 64 (11.7%) respectively. Majority 475 (79.7%) of the respondents had witnessed a seizure in the past but only 151(25.3%) and 46(7.7%) had a friend and a family member with epilepsy respectively. Among our study participants, all 596(100%) have health facility near to their home and the majority 483(81%) takes less than 30 minutes whereas 113(19%) takes 31-60 minutes.

### 3.3 Magnitude of knowledge/ attitude towards epilepsy

#### 3.3.1 Magnitude of poor knowledge

Among our study participants near to half 43.6 (95% CI: 39.6, 47.5) had poor knowledge about epilepsy. This study shows that 192 (32.2%) of the study participants reported seizure attacks didn’t originate from the brain, caused by evil spirit 278 (46.6%), and infection or injury can’t cause a seizure disorder 310 (52%). Among the study participants 27%, 42.3% and 39.5% of the respondents said epilepsy is contagious, traditional medicine is best for the treatment of epilepsy and wouldn’t mind their child to play with an epileptic child. Almost 17% of the study participants thought all children with convulsion have a seizure disorder. 177 (29.7%) thoughts seizure disorders occur in family, 176 (29.5%) did not know about the presence of different types of seizure disorder, 203 (34.1%) thought there is no medical test for diagnosing seizure disorder, and 216 (36.2%) reported Seizure disorder can be controlled with modern medicine.

#### 3.3.2 Magnitude of unfavorable attitude

In this study, of 596 participants near to half 41.3 (95% CI: 37.4, 45.1) had an unfavorable attitude towards epilepsy. Who didn’t agree their family members to marry epileptics accounts 126 (21.1%), didn’t agree to recruit epileptics 105 (17.6%), keep their child from contacting epileptics (95 (15.9%) and didn’t agree to live together with epileptics 91 (15.3%) (See table 3).

**Table 2.** Attitude towards people living with epilepsy.

| Attitude towards epilepsy | strongly agree (%) | Agree (%) | Disagree (%) | Strongly disagree (%) |
| --- | --- | --- | --- | --- |
| Do you agree to work with epileptics? | 141 (23.7) | 289 (48.5) | 103 (17.3) | 63 (10.6) |
| Do you agree to have close relation with epileptics? | 129 (21.6) | 271 (45.5) | 127 (21.3) | 69 (11.6) |
| Do you agree to live together with epileptics? | 114 (19.1) | 251 (42.1) | 140 (23.5) | 91 (15.3) |
| Do you think that epileptics shouldn't be isolated from the community? | 280 (47) | 244 (40.9) | 53 (8.9) | 19 (3.2) |
| Do you think epileptics can manage their family? | 142 (23.8) | 304 (51) | 100 (16.8) | 50 (8.4) |
| Do you shake hands of epileptics? | 251 (42.1) | 274 (46) | 36 (6) | 35 (5.9) |
| Do you keep your child from contacting epileptics? | 120 (20.1) | 244 (40.9) | 137 (23) | 95 (15.9) |
| Do you agree to recruit epileptics as a servant? | 74 (12.4) | 182 (30.4) | 235 (39.4) | 105(17.6) |
| Do you agree with your family member marry epileptics? | 72 (12.1) | 177 (29.7) | 221 (37.1) | 126 (21.1) |
| Is Epilepsy treatable disease? | 168 (28.2) | 290 (48.7) | 84 (14.1) | 54 (9.1) |
| Epileptics should not learn in schools? | 259 (43.5) | 195 (32.7) | 77 (12.9) | 65 (10.9) |
| Do you think epileptics can lead a healthy lifestyle? | 154 (25.8) | 273 (45.8) | 125 (21) | 44 (7.4) |

**Table 3.** Associated factors of poor knowledge towards epilepsy.

| Variable |  | knowledge |  | COR(95%CI) | AOR(95%CI) |
| --- | --- | --- | --- | --- | --- |
|  |  | Good | Poor |  |  |
| Age | 18-35 | 234 | 138 | 1 | 1 |
|  | 35-45 | 67 | 63 | 1.59 (1.06, 2.39)* | 0.95 (0.58, 1.57) |
|  | >45 | 35 | 59 | 2.86 (1.79, 4.56)* | 1.37 (0.74, 2.53) |
| Income | <3000 | 37 | 46 | 1 | 1 |
|  | >=3000 | 299 | 214 | 0.58 (0.36, 0.92)* | 0.78 (0.45, 1.37) |
| Occupation | Farmer or housewife | 30 | 68 | 4.47 (2.66, 7.51)* | 1.39 (0.69, 2.79) |
|  | Private workers | 95 | 75 | 1.56 (1.02, 2.37)* | 1.07 (0.64, 1.79) |
|  | Daily laborer | 34 | 27 | 1.56 (0.87, 2.80) | 0.77 (0.36, 1.64) |
|  | Student | 43 | 22 | 1.01 (0.56, 1.82) | 0.92 (0.41, 2.06) |
|  | Gov't employee | 134 | 68 | 1 | 1 |
| Educational status | Not attend modern education | 32 | 84 | 6.92 (4.21, 11.36)* | **3.74 (1.80, 7.70)\*\*** |
|  | 1-8 | 52 | 43 | 2.18 (1.33, 3.57)* | 1.45 (0.78, 2.69) |
|  | 9-12 | 78 | 67 | 2.26 (1.47, 3.49)* | **2.13 (1.27, 3.59)\*** |
|  | >12 | 174 | 66 | 1 | 1 |
| Marital status | Single | 178 | 85 | 1 | 1 |
|  | Married | 158 | 175 | 2.32 (1.66, 3.24)* | **1.88 (1.20, 3.21)\*** |
| Family role | Family head | 192 | 194 | 1 | 1 |
|  | Son or daughter | 57 | 34 | 0.59 (0.37, 0.94)* | 1.45 (0.67, 3.15) |
|  | Live alone | 87 | 32 | 0.36 (0.23, 0.57)* | 0.70 (0.37, 1.32) |
| Witnessed a seizure | Yes | 291 | 184 | 1 | 1 |
|  | No | 45 | 76 | 2.67 (1.77, 4.03)* | **1.79 (1.10, 2.90)\*** |
| Heard epilepsy | Yes | 322 | 224 | 1 | 1 |
|  | No | 14 | 36 | 3.70 (1.95, 7.01)** | **2.67 (1.27, 5.59)\*** |

Key: \*P-value < 0.05, \*\* P-value = 0.00

### 3.4 Associated factors of knowledge and Attitude towards epilepsy

#### 3.4.1 Associated factors of poor knowledge

To determine factors associated with the knowledge about epilepsy, binary logistic regression was done and computed with each independent variable. In Binary logistic regression, respondent’s age, marital status, educational status, occupation, income, family role, witnessed a seizure and heard the term epilepsy in the past were the variables that showed statistically significant association with the outcome variable that is knowledge about epilepsy. In multivariate analysis; educational status, marital status, witnessed a seizure, and heard the term epilepsy in the past was the variables that showed statistically significant association with the outcome variable that is knowledge about epilepsy. Respondents who didn’t attend modern education and those who completed secondary education were around 4 times (AOR=3.74, 95% CI: 1.80, 7.70) and 2 times (AOR=2.13, 95% CI: 1.27, 3.59) more likely to have poor knowledge than those who completed grade 12 respectively. Those participants who were married were 2 times more likely to have poor knowledge (AOR=1.88, 95% CI: 1.20, 3.21) than those who were single. Those who didn’t witness seizure were 2 times more likely to have poor knowledge (AOR=1.79, 95% CI: 1.10, 2.90) than those who witnessed seizure in the past. A respondent who hadn’t heard the term epilepsy through different mass media or training was around 3 times more likely to have poor knowledge (AOR=2.67 95% CI: 1.27, 5.59) than those who had heard about epilepsy (see table 4).

**Table 4.**
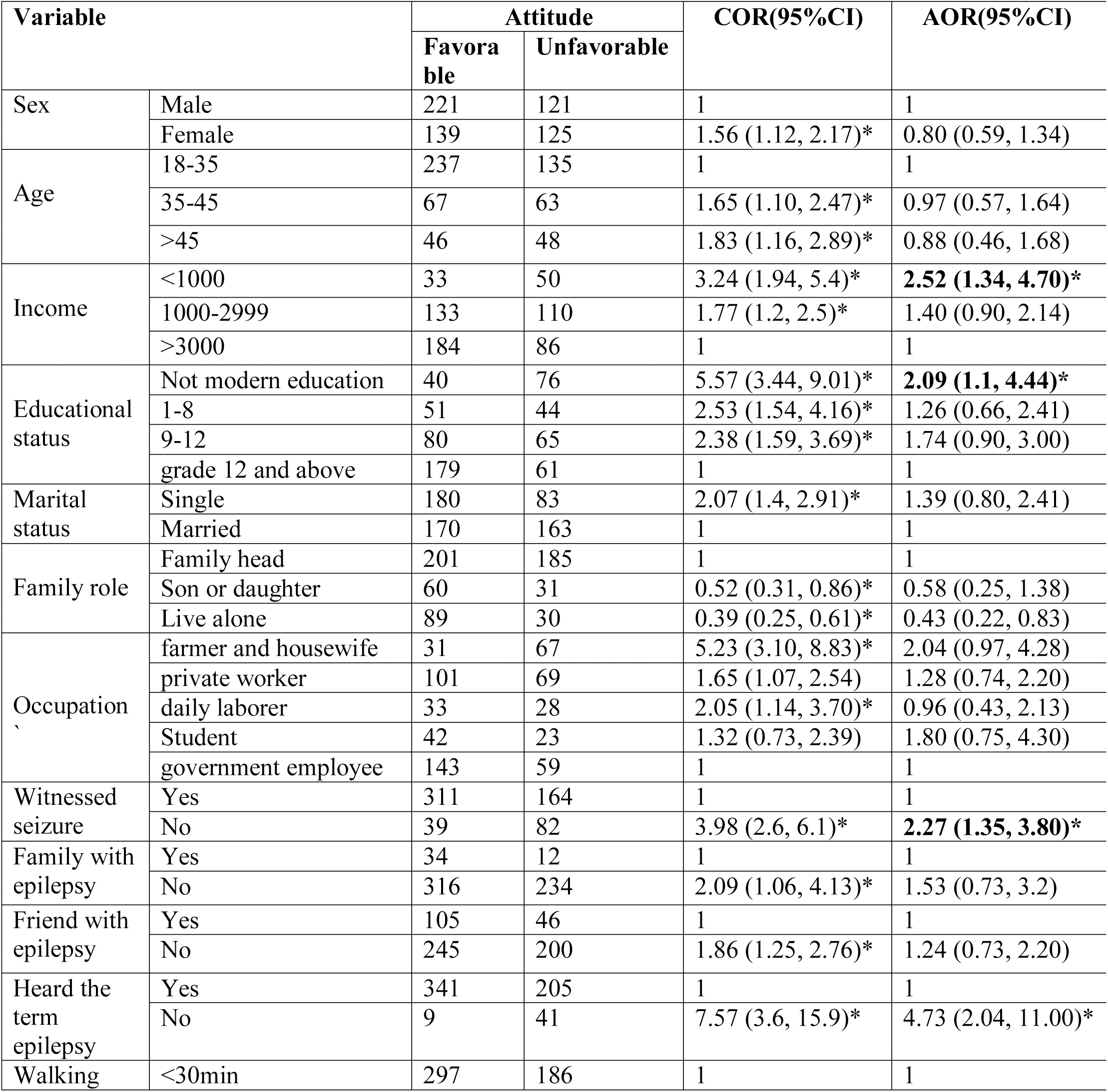
Associated factors of Attitude towards people living with epilepsy

#### 3.4.2 Associated factors of unfavorable Attitude towards people living with epilepsy

The following factors were found to be significantly associated with the attitude towards epilepsy in bivariate analysis respondent’s sex, age, monthly income, educational status, marital status, family role, occupation, whether they had witnessed seizure, whether they had a family member with epilepsy, whether they had a friend with epilepsy, whether they had heard about epilepsy and the distance of their home from a health institution.

In multivariate analysis; educational status, average monthly income, walking time from home to the health institution, witnessed a seizure, and heard the term epilepsy in the past was the variables that showed statistically significant association with the outcome variable that is knowledge about epilepsy. Respondents who earn less than 1000 Ethiopian Birr (ETB) were 2.5 times higher in odds to have unfavorable attitude than those respondents who earn average monthly income greater than 3000, (AOR=2.52, 95%CI: 1.34, 4.7). Those respondents with who were not attending modern education were 2 times more likely to have an unfavorable attitude (AOR=2.1, 95%CI: 1.10, 4.44) than those who completed grade 12.

There was also a significant association between level of attitude and whether the respondent has witnessed seizure or not; those respondent who was not witnessed seizure were 2 times likely to have unfavorable attitude than those who had witnessed a seizure, (AOR= 2.27, 95%CI: 1.35, 3.80). A respondent who did not hear the term epilepsy through different mass media or training were 5 times more likely to have unfavorable attitude than those who had heard information about epilepsy, (AOR=4.73 95%CI: 2.04, 11.00). Further, those respondents who lived far from health institutions (>30 minutes on foot walk) were 2 times more likely to have unfavorable attitude than respondents who lived in near to health institution (AOR=1.77, 95%CI: 1.06, 2.79) (see table 5).

## 4. Discussion

According to the result of this study, 43.6% (95% CI: 39.6, 47.5) of the respondent had poor general knowledge and 41.3% (95%CI: 37.4, 45.1) had an unfavorable attitude towards epilepsy. Being secondary education, marital status witnessed a seizure and heard the term epilepsy were showed statistically significant association with poor knowledge about epilepsy. Level of education, low average monthly income, not witnessed a seizure, not heard the term epilepsy and distant from health facility showed statically significant association with the unfavorable attitude.

The overall poor knowledge magnitude was in line with the study conducted in Ethiopia (20, 23). But it is much lower than studies done in southwest Ethiopia (21) and united states (19). This might be due to the study year. The study from the United States was done in 2003 and the study done in Ethiopia was during 2015.

Regarding the attitude towards epilepsy in our study, 41.3 (95%CI: 37.4, 45.1) of the respondent had an unfavorable attitude towards epilepsy. This study result is in line with the study done in south India (11).but, this finding is lower than the study conducted in Ethiopia(21, 23), Uganda (24), Nigeria (25) and Pakistan (26). This might be a difference in community belief and literacy rate across the countries.

But, the current study results are higher than the study conducted in south Indian (11); this may be due to in this study area more than half of the respondents had experience about epilepsy so those individuals may have a favorable attitude. Negative attitudes appeared to be reinforced by beliefs that epilepsy is a kind of insanity.

This study shows that respondents who didn’t attend modern education and those who were primary/secondary education were around 4 times and 2 times more likely to have poor knowledge than those who completed grade 12; this finding is in line with the study conducted in Ethiopia, the United States and Ghana (11, 23, 27). The possible justification may be individuals in higher education may expose more to epilepsy awareness creation sessions. In fact, it is expected to have knowledge change while academic status increased.

The result reveals respondents who hadn’t heard the term epilepsy through different mass media or training were around 3 times more likely to have poor knowledge than those who had heard information about epilepsy; this finding is in line with the study conducted in Ethiopia (23). Hearing the term epilepsy may invite individuals to ask/read about epilepsy and this allows to have good knowledge.

This study shows that those respondents who did not hear the term epilepsy through different mass media or training were 5 times more likely to have unfavorable attitude than those who had heard information about epilepsy; this result is in line with the study conducted in Ethiopia, which shows respondents who had heard information about epilepsy had favorable attitude compared with those who had not had information about epilepsy(23). This might be due to mass Media and training are the main strategies to change people’s attitude towards persons with epilepsy.

Study participants who earn less than 1000 Ethiopian birr were more likely to have unfavorable attitude than who earn greater than 3000 Ethiopian birrs per month. The possible reason might be due to having more monthly income associated with higher in participants’ educational level and this implies, they became employed. Living far from the health institution was also a contributor for having an unfavorable attitude and this might be due to decreased exposure level for health issues like epilepsy.

## 5. Conclusions

In this study, Debre Berhan communities were found to have deficits in terms of general knowledge and attitude about epilepsy. The knowledge and attitude towards epilepsy are likely to have an important impact on stigmatization and treatment-seeking behavior, and it should be given due attention. Lower educational status, those who didn’t witness seizure and hadn’t heard information were related to poor knowledge. Low level of income, those who didn’t witness a seizure, hadn’t heard information, lower educational status and those who lived far from health institution were related to unfavorable attitude

## 6. Recommendation

### 6.1. To the health office

The authors recommend that education program aimed at improving knowledge and attitude towards epilepsy should get attention in the study area

### 6.2. To Care provider

The care provider should give health information dissemination for the client and attendant that promote knowledge and attitude towards epilepsy during OPD visits.

## 7. Strength and limitation of the study

### 7.1. Strength

Though, the questionnaire filled in front of the participants due explanations were provided to the participants that only their spontaneous responses were required and that there were no right or wrong choices amongst the options provided for each questionnaire item.

### 7.2. Limitation

This study is a cross-sectional study and therefore the associations between variables described may not necessarily be causal or explain the change of knowledge and attitude over time in the source population. Some questionnaires were completed in front of the interviewers.

## Data Availability

All relevant materials and data supporting the findings of this study are contained within the manuscript. The remaining data will be available with a reasonable request of the corresponding author.

## 8. Declarations

### 8.2. Ethics approval

Ethical clearance was obtained from Debre Berhan University research ethics committee and an official letter of cooperation was written from department of public health to Debre Berhan town Kebele 07 and 09 administration office to obtain their consent about the reason of study and its procedure. Oral consent was obtained from study participant and the study participant was informed about issues of confidentiality and that they have right not to respond to the questions if they didn’t want to respond or to terminate the interview at any time. Each study participant was adequately informed about the objective of the study. Data was collected from all community members who had given their consent to participate in the study at the selected Kebele.

### 8.3. Availability of data

All relevant materials and data supporting the findings of this study are contained within the manuscript. The datasets used and/or analyzed during the current study are available from the corresponding author on reasonable request.

### 8.4. Authors’ contributions

AD, NA, and EA contributed to the design of the study, analysis, interpretation, and write up the manuscript. AD critically revised the final write up and analysis of the manuscript. All authors approved the final manuscript.

### 8.6. Consent for publication

“Not applicable”.

## 8.8. Acknowledgment

It is our pleasure to acknowledge Debre Berhan University for writing letter cooperation for each study Kebele and the study participants for their voluntariness.

